# Prevalence and early-life determinants of mid-life multimorbidity: evidence from the 1970 British birth cohort

**DOI:** 10.1101/2020.06.09.20126235

**Authors:** Dawid Gondek, David Bann, Matt Brown, Mark Hamer, Alice Sullivan, George B. Ploubidis

## Abstract

**Objectives:** We sought to: (1) estimate the prevalence of multimorbidity at age 46-48 in the 1970 British Cohort Study—a nationally representative sample in mid-life; and (2) examine the association between early-life characteristics and mid-life multimorbidity in the 1970 British Cohort Study.

**Design:** Prospective longitudinal birth cohort.

**Setting:** A community based sample from the 1970 British Cohort Study (BCS70).

**Participants:** All surviving children born in mainland Britain in a single week in April 1970; the analytical sample was those with valid data at age 46-48 (n=7,951; 2016-2018).

**Main outcome measure:** Multimorbidity was operationalised as a binary indicator of two or more long-term health conditions where at least one of these conditions was of physical health. It also included symptom complexes (e.g. chronic pain), sensory impairments, and alcohol problems.

**Results:** Prevalence of mid-life multimorbidity was 33.8% at age 46-48. Those with fathers from unskilled social occupational class (vs. professional) at birth had 43% higher risk of mid-life multimorbidity (risk ratio=1.43, 95% confidence interval 1.15 to 1.70). After accounting for a range of potential child and family confounders, an additional kilogram of birthweight was associated with 10% reduced risk of multimorbidity (risk ratio=0.90, 95% confidence interval 0.84 to 0.96); a decrease of one body mass index point at age 10 was associated with 3% lower risk (risk ratio=1.03, 95% confidence interval 1.01 to 1.05); one standard deviation higher cognitive ability score at age 10 corresponded to 4% lower risk (risk ratio=0.96, 95% confidence interval 0.91 to 1.00); an increase of one internalising problem at age 16 was equated with 4% higher risk (risk ratio=1.04, 95% confidence interval 1.00 to 1.08) and of one externalising problem at age 16 with 6% higher risk (risk ratio=1.06, 1.03 to 1.09).

**Conclusion:** Prevalence of multimorbidity was high in mid-life (33.8% at age 46-48) in Britain, with those in a more disadvantaged social class a birth being disproportionally affected. Potentially modifiable early-life exposures including early-life social circumstances, cognitive, physical and emotional development were associated with mid-life multimorbidity.

**What is already known on this topic?:**

- Due to differences in outcome definition, estimates of multimorbidity prevalence in mid-life (age 40-60) have varied extensively in high-income countries—from 15 to 80% between 1961 and 2013.
- There is a lack of contemporary national data in Great Britain describing the burden and nature of multimorbidity according to an agreed definition.
- The association between early-life risk factors and individual health conditions have been widely studied, however it is unknown if they are associated with multimorbidity.

**What this study adds:**

- Prevalence of multimorbidity in mid-life (age 46-48) was 33.8% in a nationally representative birth cohort in 2016-2018.
- Disadvantaged early-life parental social class, lower birthweight, lower cognitive ability, higher childhood body-mass index, and a higher number of internalising and externalising problems were found to be associated with a higher mid-life multimorbidity.

## Introduction

The prevalence of Multimorbidity has increased over the last two decades in high income countries and such trends are projected to continue.^1-3^ This presents a challenge to quality of life, due to increased risks related to polypharmacy and complex health needs among those with multimorbidity.^3^ Yet, research on multimorbidity is still relatively sparse—particularly among middle-aged individuals, who will constitute the future older population.^4^ Due to the use of various definitions, the estimates of the prevalence of multimorbidity among middle-age individuals (age 40-60) has ranged widely in high-income countries: from around 15% to 80% in the period of 1961-2013.^5^ Hence, the Academy of Medical Sciences recommended in their international policy report—published in 2018 to facilitate public health policies and interventions— to estimate the burden and nature of multimorbidity using an agreed definition as well as studying modifiable determinants of common clusters of diseases.^4^

In this study, we focused on early-life determinants of multimorbidity, as they may set a life-course trajectory of adverse exposures—predisposing to adult morbidity, hence arguably being the most appropriate life-phase for preventative efforts.^6 7^ There is also a methodological advantage of studying early-life exposures as reverse causality is less likely and they cannot be confounded by subsequent adult characteristics. We identified potentially modifiable early-life characteristics across multiple domains of early-life development, which have been studied in the context of adult morbidity outcomes separately, yet few studies have examined associations with multimorbidity. These are birthweight, social class at birth, cognitive ability and body-mass index (BMI) in childhood, and internalising and externalising problems in adolescence.

Lower birthweight^8^ and lower social class at birth^9^ have been linked with increased multimorbidity risk, yet existing evidence is limited to regional cohorts (the Hertfordshire Cohort Study in England and the Aberdeen Children of the 1950s in Scotland respectively). The association between higher BMI and adult multimorbidity risk has, to our knowledge, only been investigated cross-sectionally,^10 11^ despite extensive literature on its links with other health outcomes.^12^ Other limitations of previous studies, which this research aims to address, include not using a standardised definition of multimorbidity,^8-11 13^ not accounting for a wide range of confounders—particularly family characteristics,^9-11 13^ and focusing on specific risk factors^9-11^ as opposed to ones ranging across various domains. We have not identified any studies on the association between childhood cognitive ability and multimorbidity, despite its link with long-term sickness in mid-life, which was found to be independent of adult social class.^14^ Childhood emotional development, defined as internalising and externalising problems, has also not been studied in the context of multimorbidity. However, longitudinal studies conducted in the United Kingdom have found an association between negative affect, aggression as well as anxiety at age 13-15 with somatic and psychiatric symptoms at age 43.^13^

The 1970 British Cohort Study (BCS70) is a prospective representative cohort of those born in 1970 in Great Britain, and it comprises a rich array of information collected from birth until mid-life (age 46-48) including recently collected (in 2016-2018) biomedical as well as self-reported data.^15^ Hence, this dataset is well suited to address the objectives of our study. These are: (1) to estimate the prevalence of multimorbidity in mid-life (age 46-48) in the 1970 British Cohort Study (BCS70); and (2) to examine the association between early-life characteristics and mid-life multimorbidity as well as its components. We hypothesised that multiple early-life characteristics would be associated with a higher mid-life multimorbidity risk—lower birthweight, a more disadvantaged social class at birth, higher BMI, lower cognitive ability, and a greater number of internalising and externalising problems.

## Methods

### Participants

The history, design, and features of the BCS70 have been described.^16 17^ In brief, the survey includes all surviving children born in England, Scotland and Wales in a single week in April 1970 (n=17,196).^16^ Our analytical sample included those who participated in the data sweep at age 46-48 (from 2016-2018; n=7,951). The aim of this survey was to collect key information on cohort members’ socio-economic circumstances and health, with a range of bio-measures administered by a nurse (e.g. anthropometric and blood pressure measurements).^15^

### Measures

#### Multimorbidity

Multimorbidity was operationalised according to the definition recommended by the National Institute for Health and Care Excellence (NICE): presence of “two or more long-term health conditions where at least one of these conditions must be a physical health condition”.^18; p. 17^ These can include physical and mental health conditions, symptom complexes (e.g. chronic pain), sensory impairment, alcohol and substance misuse.^18^ Multimorbidity comprised self-reported conditions diagnosed since the previous interview (four years or more) (e.g. asthma, heart problems; see supplemental table 1 for the full list), alcohol problems (Alcohol use disorders identification test; primary care ≥5)^19^, mental health problems (Malaise Inventory ≥ 4)^20^, hypertension (systolic blood pressure ≥140 mmHg or diastolic blood pressure ≥ 90 mmHg or taking medications), and diabetes (Glycated Haemoglobin of ≥48 mmol/mol (≥ 6.5%) or taking medications; see supplemental table 2).

#### Exposures

Birthweight (kg) was recorded in the birth survey by a midwife who attended the delivery. BMI at age 10 was derived from a measure of weight and height obtained by a range of different health practitioners. Father’s social class at birth (SES) refers to the occupation of the father coded according to the Registrar General’s classification (I – professional, II – managerial and technical, III – skilled non-manual/manual, IV – partly-skilled, and V – unskilled).^21^ Cognitive ability was assessed by a modified version of the British Ability Scales^22^ at age 10. Following the approach used in previous studies, we performed a principal components analysis for each of the verbal and nonverbal sub-tests, in order to obtain scores indicating a general cognitive ability factor (g).^23 24^ The scores were standardised to a mean of zero and a standard deviation of one. Internalising and externalising problems were captured with the modified version of the Rutter A scale at age 16, completed by mothers of the participants as part of the home interview.^25^ See supplemental table 3 for details on measures of exposures.

#### Potential confounders

We control for a range of child and family characteristics which to the literature suggests may confound the association between early-life exposures and adult health (see supplemental table 4 for details on measures of confounders and supplemental table 5 for the list of studies accounting for the confounders). These include gestational age, birthweight and father’s social class at birth—when they were not used as exposures, maternal smoking during pregnancy, breastfeeding, mother’s height, mother’s marital status at birth, mother being a teen at birth, household tenure (age 5-10), overcrowding (>1 person per room at age 5), teacher’s report of parental interest in child’s education (age 5-10), length of time absent from school due to illness (age 10) and parental divorce (age 16).

### Missing data

Multimorbidity was deemed missing if participants had incomplete information on at least one component of multimorbidity (n=3,793 out of 7,951 defined as the study sample). To restore sample representativeness and reduce selection bias, we used multiple imputation with chained equations generating 50 datasets.^26^ In order to investigate sensitivity of the estimates due to missing information, the prevalence of multimorbidity is also presented under different missing data generating mechanisms and across imputations based on samples with varying missing data inclusion criteria (see supplemental text 1 and supplemental table 6 for more details).

### Analysis

#### Exposures—multimorbidity association

The associations between exposures (birthweight, father’s social class at birth, cognitive ability at age 10, BMI at age 10, externalising as well as internalising problems at age 16) and multimorbidity (age 46-48) in BCS70 were estimated with multivariate Poisson regression. We present gender-adjusted estimates and further adjusted models that account for a range of potential child and family confounders (i.e., common causes of both exposures and multimorbidity).

As a robustness check, the analysis was also re-run using a count of health conditions as an outcome. We also tested—using the Wald test—for non-linear associations between continuous exposures and multimorbidity by including squared and linear terms (e.g. birthweight^2^ and birthweight) in unadjusted regression models, but there was no evidence of departure from linearity. Likewise, using a similar approach no evidence of gender*exposure interaction was found.

#### Exploratory analysis – multimorbidity as clusters of conditions

Due to heterogeneous definitions of multimorbidity,^4 27^ we additionally investigated the determinants of five most common pairs of conditions (and their individual components): mental health morbidity and hypertension, mental health morbidity and arthritis, mental health and diabetes, mental health morbidity and asthma/bronchitis, diabetes and hypertension.^28^ As the clusters used in these analyses were derived from the multimorbidity outcome, they are likely to be closely-related hence increasing family-wise error rate. Thus, we employ a more stringent p-value threshold of 0.003 —using the Bonferroni correction (α = 0.05 divided by 20 tests).^29^ We evaluate our findings with this threshold, but also consider the strength of association, coverage of confidence intervals and pattern of p-values.

## Results

### Prevalence of multimorbidity

The prevalence of multimorbidity in BCS70 at age 46-48 was 33.8%. The most prevalent individual health outcomes were high-risk drinking (26.3%), recurrent back problems (20.9%), and mental health problems (19.1%) (see table 1). Among the most prevalent chronic physical health conditions were asthma/bronchitis (11.7%) and arthritis (7.7%).

**Table 1.**
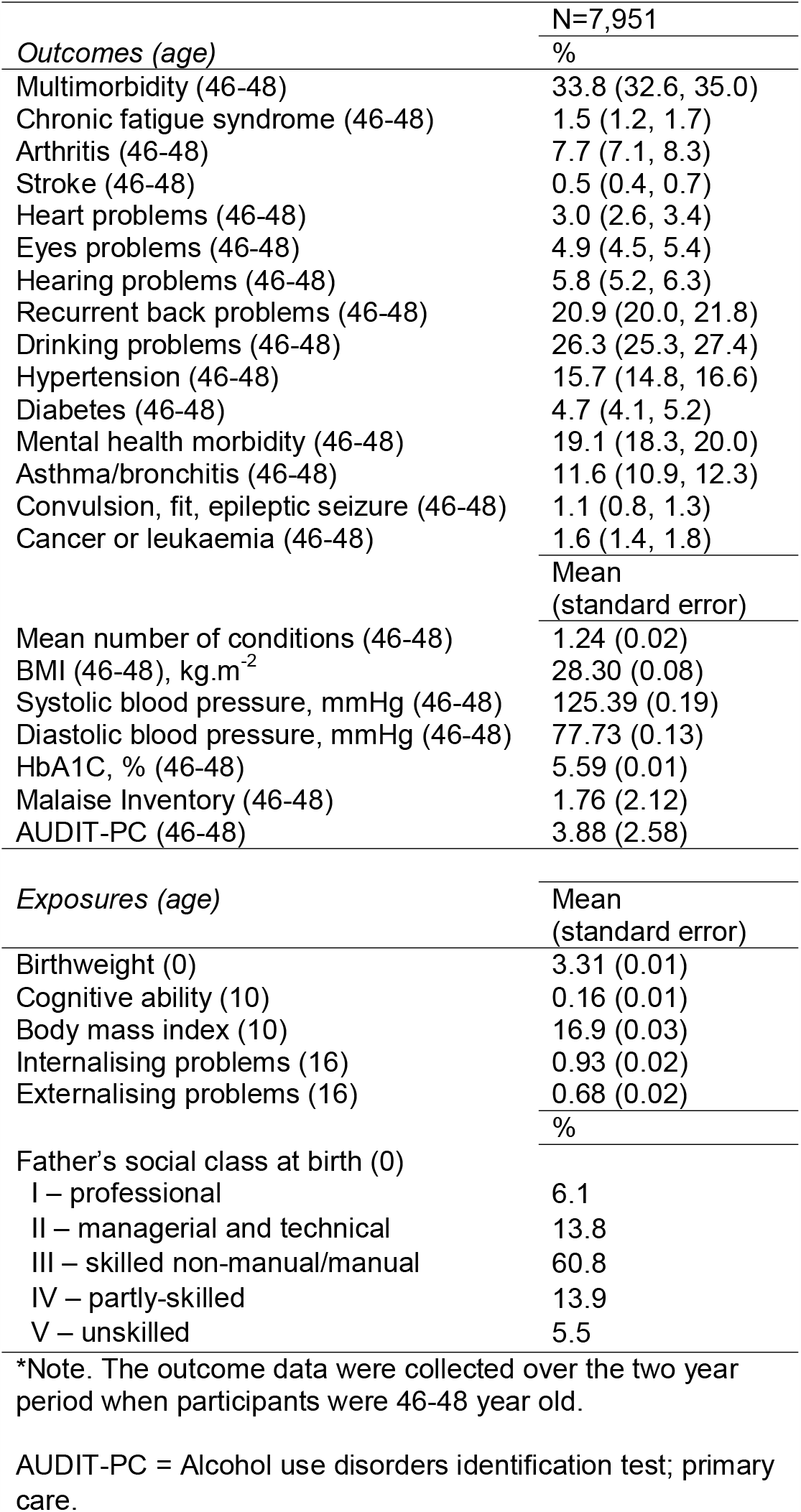
Descriptive statistics of the health outcomes and early-life exposures in the 1970 British birth cohort (BCS70).

|  | N=7,951 |
| --- | --- |
| *Outcomes (age)* | % |
| Multimorbidity (46-48) | 33.8 (32.6, 35.0) |
| Chronic fatigue syndrome (46-48) | 1.5 (1.2, 1.7) |
| Arthritis (46-48) | 7.7 (7.1, 8.3) |
| Stroke (46-48) | 0.5 (0.4, 0.7) |
| Heart problems (46-48) | 3.0 (2.6, 3.4) |
| Eyes problems (46-48) | 4.9 (4.5, 5.4) |
| Hearing problems (46-48) | 5.8 (5.2, 6.3) |
| Recurrent back problems (46-48) | 20.9 (20.0, 21.8) |
| Drinking problems (46-48) | 26.3 (25.3, 27.4) |
| Hypertension (46-48) | 15.7 (14.8, 16.6) |
| Diabetes (46-48) | 4.7 (4.1, 5.2) |
| Mental health morbidity (46-48) | 19.1 (18.3, 20.0) |
| Asthma/bronchitis (46-48) | 11.6 (10.9, 12.3) |
| Convulsion, fit, epileptic seizure (46-48) | 1.1 (0.8, 1.3) |
| Cancer or leukaemia (46-48) | 1.6 (1.4, 1.8) |
|  | Mean<br>(standard error) |
| Mean number of conditions (46-48) | 1.24 (0.02) |
| BMI (46-48), kg.m<sup>-2</sup> | 28.30 (0.08) |
| Systolic blood pressure, mmHg (46-48) | 125.39 (0.19) |
| Diastolic blood pressure, mmHg (46-48) | 77.73 (0.13) |
| HbA1C, % (46-48) | 5.59 (0.01) |
| Malaise Inventory (46-48) | 1.76 (2.12) |
| AUDIT-PC (46-48) | 3.88 (2.58) |
| *Exposures (age)* | Mean<br>(standard error) |
| Birthweight (0) | 3.31 (0.01) |
| Cognitive ability (10) | 0.16 (0.01) |
| Body mass index (10) | 16.9 (0.03) |
| Internalising problems (16) | 0.93 (0.02) |
| Externalising problems (16) | 0.68 (0.02) |
|  | % |
| Father's social class at birth (0) |  |
| I – professional | 6.1 |
| II – managerial and technical | 13.8 |
| III – skilled non-manual/manual | 60.8 |
| IV – partly-skilled | 13.9 |
| V – unskilled | 5.5 |

\*Note. The outcome data were collected over the two year period when participants were 46-48 year old.
AUDIT-PC = Alcohol use disorders identification test; primary care.

### Exposures—mid-life multimorbidity association

In gender-adjusted models, all exposures were associated with a greater risk of multimorbidity at age 46-48: lower birthweight, lower cognitive ability at age 10, higher BMI at age 10, more internalising and externalising problems at age 16 as well as a more disadvantaged father’s social classes at birth (p<0.001): with unskilled class having 43% higher risk of multimorbidity (risk ratio; RR=1.43, 95% confidence interval 1.15 to 1.70) (table 2).

**Table 2.** Association between early-life exposures and multimorbidity at age 46-48.

| N=7,951 | Relative risk (95%CI) |  |
| --- | --- | --- |
|  | Gender-adjusted | Confounders-adjusted |
| SES at birth (age 0) |  | N/A* |
| I – professional | 1.00 | 1.00 |
| II – managerial and technical | 1.14 (0.94, 1.40) | - |
| III – skilled non-manual/manual | 1.30 (1.09, 1.55) | - |
| IV – partly-skilled | 1.43 (1.18, 1.74) | - |
| V – unskilled | 1.43 (1.15, 1.77) | - |
| Birthweight (age 0) | 0.86 (0.80, 0.91) | 0.90 (0.84, 0.96)<sup>1</sup> |
| Cognitive ability (age 10) | 0.91 (0.87, 0.95) | 0.96 (0.91, 1.00)<sup>2</sup> |
| BMI (age 10) | 1.03 (1.01, 1.05) | 1.03 (1.01, 1.05)<sup>3</sup> |
| Internalising problems (age 16) | 1.07 (1.04, 1.11) | 1.04 (1.00, 1.08)<sup>4</sup> |
| Externalising problems (age 16) | 1.09 (1.07, 1.12) | 1.06 (1.03, 1.09)<sup>5</sup> |

\*Not adjusted for any other variables as they may potentially lie on the causal pathway.
<sup>1</sup> Adjusted for gender, father's social class at birth, mother ever smoked during pregnancy, mother's height, mother's marital status at birth, mother being a teen at birth;
<sup>2</sup> Adjusted for gender, father's social class at birth, birthweight, mother ever smoked during pregnancy, mother breastfed, mother's height, mother's marital status at birth, mother being a teen at birth, household tenure (5-10), parental interest in child's education (10), overcrowding (5), parental interest in child's education (10), length of time absent from school due to illness (10), parental divorce (16), mother's mental health (10), BMI at age 10;
<sup>3</sup> Adjusted for gender, father's social class at birth, birthweight, mother ever smoked during pregnancy, mother breastfed, mother's height, mother's marital status at birth, mother being a teen at birth, household tenure (5-10), parental interest in child's education (10), overcrowding (5), parental interest in child's education (10), length of time absent from school due to illness (10), parental divorce (16), mother's mental health (10), cognitive ability at age 10, externalising and internalising problems at age 16;
<sup>4</sup> Adjusted for gender, father's social class at birth, birthweight, mother ever smoked during pregnancy, mother breastfed, mother's height, mother's marital status at birth, mother being a teen at birth, household tenure (5-10), parental interest in child's education (10), overcrowding (5), parental interest in child's education (10), length of time absent from school due to illness (10), parental divorce (16), mother's mental health (10), cognitive ability and BMI at age 10, and externalising problems at age 16;
<sup>5</sup> Adjusted for gender, father's social class at birth, birthweight, mother ever smoked during pregnancy, mother breastfed, mother's height, mother's marital status at birth, mother being a teen at birth, household tenure (5-10), parental interest in child's education (10), overcrowding (5), parental interest in child's education (10), length of time absent from school due to illness (10), parental divorce (16), mother's mental health (10) cognitive ability and BMI at age 10, and internalising problems at age 16.

Adjustment for potential confounders had little effect on the strength of the associations. An additional kilogram of birthweight was associated with 10% reduced risk of multimorbidity (RR=0.90, 0.84 to 0.96); a decrease of one point on BMI scale was associated with 3% lower risk (RR=1.03, 1.01 to 1.05); one standard deviation higher score on cognitive ability measure corresponded to 4% lower risk (RR=0.96, 95% CI 0.91 to 1.00); increase of one internalising problem was equated with 4% higher risk (RR=1.04, 1.00 to 1.08) and of one externalising problem with 6% higher risk (RR=1.06, 1.03 to 1.09).

### Exploratory analysis—multimorbidity as clusters of conditions

The prevalence of pairs of conditions were: mental health (MH)/hypertension (4.1%), MH/asthma (3.3%), MH/arthritis (2.5%), diabetes/hypertension (2.1%), MH/diabetes (1.4%).

As presented in table 3, there was strong evidence (at the Bonferroni corrected p<0.003) for the association between father’s SES at birth and clusters including mental health problems: MH/hypertension (for unskilled vs professional class: RR=2.92, 1.23 to 6.94) and MH/arthritis (RR=3.41, 1.46 to 7.95). These associations were somewhat stronger than the ones found for the individual conditions, as shown in table 4: arthritis (for unskilled vs professional class: (RR=1.86, 1.19 to 2.91), hypertension (RR=1.52, 1.05 to 2.20), and mental health problems (RR=1.54, 1.11 to 2.10; with p-value being slightly above the Bonferroni threshold: p=0.003). However, the association between father’s SES and diabetes was also particularly strong (e.g. for unskilled vs professional class: RR=3.29, 1.26 to 8.56).

**Table 3.** The association between early-life risk factors and multimorbidity clusters at age 46-48.

| N=7,951 | MH + hypertension | MH + arthritis | MH + diabetes | MH + asthma | Diabetes + hypertension |
| --- | --- | --- | --- | --- | --- |
|  | Relative risk (95%CI) |  |  |  |  |
| Father's SES at birth<sup>1</sup> |  |  |  |  |  |
| I – professional (reference) | 1.00* | 1.00* | 1.00 | 1.00 | 1.00 |
| II – managerial and technical | 1.27 (0.53, 3.04) | 1.19 (0.51, 2.80) | 2.69 (0.35, 20.83) | 1.67 (0.78, 3.56) | 2.04 (0.52, 7.99) |
| III – skilled non-manual/manual | 2.32 (1.11, 4.87) | 1.53 (0.72, 3.26) | 4.40 (0.67, 29.13) | 1.68 (0.83, 3.40) | 3.01 (0.85, 10.61) |
| IV – partly-skilled | 2.96 (1.36, 6.44) | 1.92 (0.85, 4.35) | 5.13 (0.56, 30.32) | 2.03 (0.97, 4.26) | 3.59 (0.97, 13.35) |
| V – unskilled | 2.92 (1.23, 6.94) | 3.41 (1.46, 7.95) | 5.92 (0.78, 44.64) | 2.04 (0.85, 4.89) | 3.60 (0.92, 14.09) |
| Birthweight<sup>2</sup> | 0.83 (0.65, 1.05) | 0.84 (0.64, 1.10) | 1.03 (0.66, 1.61) | 0.83 (0.65, 1.07) | 0.72 (0.49, 1.06) |
| Cognitive ability (age 10)<sup>3</sup> | 0.85 (0.72, 1.00) | 0.75 (0.63, 0.89)* | 0.80 (0.61, 1.06) | 0.85 (0.72, 1.00) | 0.91 (0.73, 1.12) |
| BMI (age 10)<sup>4</sup> | 1.11 (1.05, 1.18)* | 1.03 (0.96, 1.12) | 1.16 (1.06, 1.28)* | 1.04 (0.98, 1.11) | 1.25 (1.16, 1.34)* |
| Internalising problems (age 16)<sup>5</sup> | 1.21 (1.09, 1.34)* | 1.17 (1.03, 1.33)* | 1.09 (0.87, 1.38) | 1.25 (1.12, 1.39)* | 0.97 (0.79, 1.19) |
| Externalising problems (age 16)<sup>6</sup> | 1.00 (0.90, 1.11) | 1.06 (0.95, 1.19) | 1.04 (0.87, 1.24) | 1.02 (0.91, 1.14) | 1.04 (0.89, 1.22) |

<sup>1</sup> Adjusted for gender.
<sup>2</sup> Adjusted for gender, father's social class at birth, mother ever smoked during pregnancy, mother's height, mother's marital status at birth, mother being a teen at birth;
<sup>3</sup> Adjusted for gender, father's social class at birth, birthweight, mother ever smoked during pregnancy, mother breastfed, mother's height, mother's marital status at birth, mother being a teen at birth, household tenure (5-10), parental interest in child's education (10), overcrowding (5), parental interest in child's education (10), length of time absent from school due to illness (10), parental divorce (16), mother's mental health (10), BMI at age 10;
<sup>4</sup> Adjusted for gender, father's social class at birth, birthweight, mother ever smoked during pregnancy, mother breastfed, mother's height, mother's marital status at birth, mother being a teen at birth, household tenure (5-10), parental interest in child's education (10), overcrowding (5), parental interest in child's education (10), length of time absent from school due to illness (10), parental divorce (16), mother's mental health (10), cognitive ability at age 10, externalising and internalising problems at age 16;
<sup>5</sup> Adjusted for gender, father's social class at birth, birthweight, mother ever smoked during pregnancy, mother breastfed, mother's height, mother's marital status at birth, mother being a teen at birth, household tenure (5-10), parental interest in child's education (10), overcrowding (5), parental interest in child's education (10), length of time absent from school due to illness (10), parental divorce (16), mother's mental health (10), cognitive ability and BMI at age 10, and externalising problems at age 16;
<sup>6</sup> Adjusted for gender, father's social class at birth, birthweight, mother ever smoked during pregnancy, mother breastfed, mother's height, mother's marital status at birth, mother being a teen at birth, household tenure (5-10), parental interest in child's education (10), overcrowding (5), parental interest in child's education (10), length of time absent from school due to illness (10), parental divorce (16), mother's mental health (10) cognitive ability and BMI at age 10, and internalising problems at age 16.
\*significant at p-value (after the Bonferroni correction) = 0.003 (for father's SES at birth, it refers to all categories combined)

**Table 4.** The association between early-life risk factors and individual conditions at age 46-48.

| N=7,951 | Mental health | Arthritis | Diabetes | Asthma | Hypertension |
| --- | --- | --- | --- | --- | --- |
|  | Relative risk (95%CI) |  |  |  |  |
| Father's SES at birth<sup>1</sup> |  |  |  |  |  |
| I – professional (reference) | 1.00 | 1.00 | 1.00 | 1.00 | 1.00 |
| II – managerial and technical | 1.15 (0.89, 1.50) | 0.86 (0.56, 1.36) | 2.11 (0.83, 5.36) | 1.31 (0.96, 1.80) | 1.05 (0.75, 1.48) |
| III – skilled non-manual/manual | 1.28 (1.01, 1.61) | 1.29 (0.89, 1.86) | 2.94 (1.22, 7.10) | 1.16 (0.87, 1.54) | 1.41 (1.05, 1.88) |
| IV – partly-skilled | 1.47 (1.14, 1.89) | 1.25 (0.83, 1.89) | 3.38 (1.37, 8.35) | 1.15 (0.83, 1.58) | 1.66 (1.22, 2.27) |
| V – unskilled | 1.54 (1.14, 2.10) | 1.86 (1.19, 2.91) | 3.29 (1.26, 8.56) | 1.09 (0.73, 1.63) | 1.52 (1.05, 2.20) |
| Birthweight<sup>2</sup> | 0.96 (0.87, 1.06) | 0.95 (0.81, 1.10) | 0.71 (0.55, 0.91) | 0.85 (0.75, 0.96) | 0.80 (0.72, 0.89)* |
| Cognitive ability (age 10)<sup>3</sup> | 0.90 (0.84, 0.96)* | 0.92 (0.84, 1.02) | 0.93 (0.79, 1.08) | 0.98 (0.90, 1.06) | 0.93 (0.87, 1.00) |
| BMI (age 10)<sup>4</sup> | 1.02 (1.00, 1.05) | 1.03 (0.99, 1.07) | 1.19 (1.13, 1.25)* | 1.00 (0.97, 1.04) | 1.10 (1.06, 1.13)* |
| Internalising problems (age 16)<sup>5</sup> | 1.18 (1.13, 1.24)* | 1.06 (0.97, 1.15) | 0.97 (0.85, 1.12) | 1.05 (0.98, 1.12) | 1.02 (0.95, 1.09) |
| Externalising problems (age 16)<sup>6</sup> | 1.01 (0.97, 1.06) | 1.08 (1.00, 1.15) | 1.07 (0.95, 1.20) | 1.03 (0.96, 1.10) | 1.02 (0.96, 1.08) |

<sup>1</sup> Adjusted for gender.
<sup>2</sup> Adjusted for gender, father's social class at birth, mother ever smoked during pregnancy, mother's height, mother's marital status at birth, mother being a teen at birth;
<sup>3</sup> Adjusted for gender, father's social class at birth, birthweight, mother ever smoked during pregnancy, mother breastfed, mother's height, mother's marital status at birth, mother being a teen at birth, household tenure (5-10), parental interest in child's education (10), overcrowding (5), parental interest in child's education (10), length of time absent from school due to illness (10), parental divorce (16), mother's mental health (10), BMI at age 10;
<sup>4</sup> Adjusted for gender, father's social class at birth, birthweight, mother ever smoked during pregnancy, mother breastfed, mother's height, mother's marital status at birth, mother being a teen at birth, household tenure (5-10), parental interest in child's education (10), overcrowding (5), parental interest in child's education (10), length of time absent from school due to illness (10), parental divorce (16), mother's mental health (10), cognitive ability at age 10, externalising and internalising problems at age 16;
<sup>5</sup> Adjusted for gender, father's social class at birth, birthweight, mother ever smoked during pregnancy, mother breastfed, mother's height, mother's marital status at birth, mother being a teen at birth, household tenure (5-10), parental interest in child's education (10), overcrowding (5), parental interest in child's education (10), length of time absent from school due to illness (10), parental divorce (16), mother's mental health (10), cognitive ability and BMI at age 10, and externalising problems at age 16;
<sup>6</sup> Adjusted for gender, father's social class at birth, birthweight, mother ever smoked during pregnancy, mother breastfed, mother's height, mother's marital status at birth, mother being a teen at birth, household tenure (5-10), parental interest in child's education (10), overcrowding (5), parental interest in child's education (10), length of time absent from school due to illness (10), parental divorce (16), mother's mental health (10) cognitive ability and BMI at age 10, and internalising problems at age 16.
\*significant at p-value (after the Bonferroni correction) = 0.003 (for father's SES at birth, it refers to all categories combined)

Birthweight was associated with diabetes and hypertension, with 1kg higher weight being linked with 29% (RR=0.71, 0.55 to 0.91) and 20% (RR=0.80, 0.72 to 0.89) lower risk of having these conditions respectively (table 4). Cognitive ability was associated with MH/arthritis (RR=0.75, 0.63 to 0.89), and mental health problems (RR=0.90, 0.84 to 0.96) (tables 3-4). Externalising problems were not found to be linked with any cluster or individual condition. Whereas, internalising problems were linked with clusters including mental health problems: MH/hypertension, MH/arthritis, MH/asthma and with mental health problems as an individual condition (table 4). BMI at age 10 had the strongest association with diabetes/hypertension clusters (RR=1.25, 1.16 to 1.34) and it was linked with diabetes and hypertension as individual conditions and their clusters with mental health (tables 3-4).

## Discussion

### Main findings

The prevalence of multimorbidity in Britain was 33.8% at age 46-48—based on the nationally representative data of BCS70. Being in less advantaged social classes at birth, lower birthweight, lower cognitive ability, higher BMI—both at age 10, more externalising and internalising problems at age 16 were found to be associated with higher mid-life multimorbidity after a rich set of confounders were accounted for.

These associations reflected different patterns of association with the components of multimorbidity. Higher childhood BMI and lower birthweight were linked with increased risk of diabetes and hypertension; internalising problems and cognitive ability were most strongly associated with comorbidities including adult mental health problems; a more disadvantaged father’s SES was linked with diabetes and MH/hypertension as well as MH/arthritis clusters. Whereas, externalising problems had a very weak association with the individual conditions or their clusters, despite being linked with the overall multimorbidity outcome.

### Comparison with previous studies and interpretation

The prevalence of multimorbidity in our study was comparable with the most comprehensive estimate of multimorbidity in mid-life in the UK—which was 30.4% among over 1.7 million general practice patients aged 45-64 in 2007.^30^

Early-life BMI was found to be associated with multimorbidity, which is consistent with the previous literature on a range of adult morbidity outcomes as well as multimorbidity specifically.^10-12 31^ The evidence on the association between childhood BMI and diabetes and hypertension, found in this study, is consistent with finding from both traditional observational research^12^ and Mendelian randomisation studies, in which genes are employed as instrumental variables.^32^ This may be due to an increased risk of dyslipidaemia and systemic inflammation due to obesity, which may constitute a common pathway to the development of both diabetes and cardiovascular conditions.^33 34^

Externalising and internalising problems have been found to be associated with a range of measures of adult morbidity;^35-37^ and with co-existing somatic and psychiatric symptoms in mid-life.^13^ However, it appears that the association between internalising problems and multimorbidity may be driven by their links with mental health problems included in the multimorbidity definition—no associations are found when mental health morbidity was removed from the multimorbidity definition (results not shown). Interestingly, externalising problems appeared to increase the risk of overall multimorbidity to a much larger extent than risk of each major individual components. There are several potential mechanisms linking early-life mental health problems with adult multimorbidity. According to the allostatic stress model, exposure to chronic stressors may result in physiological dysregulation, which predisposes an individual to poor health.^38^ For instance, there is evidence on the association between children’s depression and worse immune functioning.^39^ In addition, internalising and particularly externalising problems may have a more indirect effect on later multimorbidity, through their link with negative health behaviours, such as smoking, physical activity, and drinking.^40-44^

The link between early-life cognitive ability and adult morbidity has been previously found,^14 37^ yet there is no existing evidence on multimorbidity. We found evidence for a modest association, where an increase of one standard deviation in cognitive ability (around 15 points on a standard general intelligence test) was associated with 4% decrease in the probability of multimorbidity. There are several potential pathways linking early-life cognitive ability with adult multimorbidity, including better self-care, indirect link via health behaviours or shared pathways with education or socioeconomic position.^45^

Father’s social class at birth was associated with multimorbidity in our study, which is consistent with the extensive literature on adult morbidity^9 46-50^ and multimorbidity specifically.^5 9^ Our research adds to this literature by showing a particularly strong association between father’s SES and mental health problems clustered with hypertension or arthritis. Previous research found that the link between early-life socioeconomic circumstances is partially mediated by cognitive ability, educational attainment, and school type.^9^ Life course theory and related findings also suggest that early-life socioeconomic position increases the risk of other adverse exposures, such as negative health behaviours or unfavourable adult socioeconomic circumstances, which have a cumulative effect on health over the lifespan.^51^

In contrast to our results, a previous study found no association between birthweight and multimorbidity.^8^ However, the effect size in our study is modest, with 10% decrease in risk corresponding to 1kg change in birthweight; or 1.6% avoided cases, assuming causality, if 20% with the lowest weight were “shifted” to the mean of other 80%. Overall, the evidence on the association between low birthweight and adult morbidity is somewhat inconsistent.^52^ It appears that birthweight may be an important exposure for diabetes and hypertension, as shown by our analysis and other observational studies.^53 54^ However, findings from Mendelian Randomization studies suggests that only the link with diabetes may be causal.^53 54^ This may be due to prenatal growth stress leading to metabolic reprogramming beginning in utero,^55 56^ according to the Barker hypothesis.^57^

### Strengths and limitations

To the best of our knowledge, this is the first study examining the association between emotional development, cognitive ability and multimorbidity. The main strength of our study is that it used a contemporary and representative sample of the mid-life population born in Britain and—contrary to the previous research— accounted for a rich set of confounders, particularly parental characteristics. However, there is always a risk of bias in the estimates based on observational studies, due to omitting potential confounders, for instance, genetic factors that may affect both early-life physical and emotional development and later health. As a sensitivity check, we estimated the E-value, which indicates the minimum strength of association that an unmeasured confounder would need to have with both the treatment and outcome to fully explain away a specific treatment–outcome association, after conditioning on the measured covariates.^58^ As shown by supplemental table 8, the association between each early-life exposure and multimorbidity at age 46-48, could be explained away by an unmeasured confounder that was associated with both the exposure and outcome by a risk ratio of at least 1.21, above and beyond the measured confounders. This value is stronger than the association between any measured exposure or confounder and the outcome in this study, but the potential for bias due to unobserved confounders cannot be fully ruled out.

Our work relied on self-reported health outcomes. However, we also included objectively measured diabetes and hypertension, which are free from biases related to self-reporting. In addition, self-reports appear to be reliable measures at least in our study, as we found strong agreement between self-reported and objectively measured hypertension and diabetes (89% and 98% respectively; with fair—0.51— and good—0.74—Cohen’s Kappa).

Another limitation, common to studies using prospective longitudinal data, is selective attrition and a large proportion of missing data. Hence, we used multiple imputation to reduce bias. Multiple imputation returns unbiased results under the missing at random (MAR) assumption, which implies that systematic differences between the missing and observed values can be explained by the observed data.^59 60^ We enriched the imputation model in order to maximise the plausibility of the MAR assumption with auxiliary variables (self-perceived general health, individual health conditions under multimorbidity outcome and smoking), which were not part of the substantive model of interest, but they were related to the probability of missingness and/or related to the incomplete outcome (see supplemental table 6). We obtained similar estimates of multimorbidity prevalence from analyses under different missing data generating mechanisms (MCAR vs MAR) and across imputations based on samples with varying missing data inclusion criteria, which provides evidence for robustness of the findings (see supplemental table 7).

### Conclusions and implications

Multimorbidity affects over one-third of people born in Britain in 1970. Co-occurring mental and physical health conditions, such as diabetes or hypertension, appear to be particularly important comorbidities to target—due to their detrimental link on overall functioning.^61-63^ Early-life socioeconomic disadvantage, high BMI, low cognitive ability and poor emotional development were all associated with a higher risk of mid-life multimorbidity and various clusters of conditions. Hence, if the presented associations reflect causal effects, reducing their impact or prevalence, through both health promotion and primary prevention, may improve various aspects of mid-life health.

## Data Availability

BCS70 data used in this study have been managed by the Centre for Longitudinal Study and are accessible via the UK data archive. The statistical code for derived variables is available from the correspondence author.

## Footnotes

DG is a corresponding author. DG, GP, DB conceived and designed the study. GP and DB supervised the study. DG performed the statistical analysis. All authors contributed to interpretation of data, revised the manuscript and approved the final version before submission. DG and GP are the guarantors and attest that all listed authors meet authorship criteria and that no others meeting the criteria have been omitted.

## Acknowledgments

Dawid Gondek would like to thank Economic and Social Research Council (ESRC) +3 PhD for the support.

## Ethical approval

The BCS70 has been granted ethical approval for each sweep from 2000 by the National Health Service (NHS) Research Ethics Committee.

## Patient consent

All participants provided written informed consent after a thorough explanation of the research procedures.

## Data sharing

The lead author (DG) affirms that this manuscript is an honest, accurate, and transparent account of the study being reported; that no important aspects of the study have been omitted; and that any discrepancies from the study as planned have been explained.

Dissemination to participants and related patient and public communities: The results of this research will be reported in newsletters for study participants, and public lectures.

## Patient involvement

This research was done without patient involvement. Patients were not invited to comment on the study design and were not consulted to develop patient relevant outcomes or interpret the results. Patients were not invited to contribute to the writing or editing of this document for readability or accuracy.

